# Classifying sex with MRI

**DOI:** 10.1101/2022.04.27.22274355

**Authors:** Matthis Ebel, Martin Lotze, Martin Domin, Nicola Neumann, Mario Stanke

## Abstract

Sex differences in the size of specific brain structures have been extensively studied but careful and reproducible statistical hypothesis testing to identify them produced overall small effect sizes and differences brains of males and females. On the other hand, multivariate statistical or machine learning methods that analyse MR images of the whole brain have reported respectable accuracies for the task of distinguishing males from females. However, most existing studies lacked a careful control for brain volume differences between sexes and, if done, their accuracy often declined to 70% or below. This raises questions on the relevance of accuracies achieved without careful control of overall volume. Also the potential applicability is uncertain insofar as the robustness of methods had rarely been tested or they suffered from poor accuracy when applied on a different cohort.

We examined how accurately sex can be classified with multivariate methods from gray matter properties of the human brain when correcting for overall brain volume. We also tested, how robust machine learning classifiers are when predicting cross-cohort, i.e. when they are used on a different cohort than they were trained on. Further, we studied how their accuracy depends on the size of the training set. MRI data was used from two population based data sets of 3308 mostly older adults from the Study of Health in Pomerania (SHIP) and 1113 mostly younger adults from the Human Connectome Project (HCP), respectively. Our new open source program BraiNN is based on a 3D convolutional neural network and was compared with a simple logistic regression approach.

When using the gold standard method of matching male and female participants for total intracranial volume, BraiNN achieved 86% accuracy when predicting sex on the same (SHIP) cohort and 73% accuracy when cross-predicting on the HCP cohort. Logistic regression achieved an accuracy >90% on the SHIP cohort, but required a large number of training examples to perform well and did not generalize well across cohorts. On the other hand, BraiNN lost less than 2% accuracy when the cohort size was reduced from 3308 to 1274.

## 1. Introduction

Brains of men and women are essentially similarly structured, exhibiting only small differences in regional cortical and subcortical volumes, cortical thickness, and white matter (Eliot et al., 2021) and belonging to the same population rather than two different ones (Joel and Fausto-Sterling, 2016; Joel, 2021). Nevertheless, large cohort studies (Lotze et al., 2019; Raznahan et al., 2011; Ritchie et al., 2018; Williams et al., 2021) and meta-analyses (Ruigrok et al., 2014; Sacher et al., 2013) confirmed sex differences in global and regional brain volumes, and even if effect sizes were small, this does not mean that these differences are meaningless (Funder and Ozer, 2019). Using multivariate statistical approaches, more subtle neuroanatomical sex differences can be uncovered (Sepehrband et al., 2018). Thereby methodological issues are crucial, such as the correction for overall brain size (Sanchis-Segura et al., 2020).

We here seek differences in the brains of males and females from the machine learning viewpoint (sometimes referred to as multivariate analysis) rather the statistical view-point. This means we primarily seek accurate predictions rather than reliable and simply interpretable differences. The addressed task is sex classification, i.e. to predict whether an MR image was taken from a man or woman. Previous studies reported that brains of males and females can be differentiated with an accuracy of 69% to 96% using voxel pattern analyses (Anderson et al., 2019; Brennan et al., 2021; Chekroud et al., 2016; Kurth et al., 2021; Feis et al., 2013).

However, a large caveat to those accuracy numbers is that to our knowledge no large study on sex classification followed the gold-standard approach to correct for sex-specific average brain size differences by matching men and women of the same overall brain volume. The study of Wang et al. (2012) did so but had a sample of only 35 women and 35 men. Luders et al. (2009) used a volume-matched data set as well, but did not predict the sex.

The men’s average total intracranial volume (TIV) is larger than the women’s, by about 12% according to Ruigrok et al. (2014). As is well-known and as we quantify below for our data sets, TIV therefore allows a fairly good discrimination between men and women. Sanchis-Segura et al. (2020) reported on their data set with a narrow age range that a sex prediction accuracy of remarkable 84% can be achieved using total intracranial volume (TIV) alone. To uncover subtle differences between brains of males and females, however, the total size difference is irrelevant, the overall brain size a nuisance variable and should be corrected for in order to find relevant and direct sex differences (Luders et al., 2009). Liu et al. (2014) argue that the popular normalization practice of dividing regional volumes by overall brain volume is not properly correcting for overall volume. They instead argue for a non-linear correction term, chose a power-law fitted to the data (’power-corrected proportions’). Many studies, such as ours, apply a non-linear spacial normalization of raw input in order to measure the volumes in a reference grid of voxels. This mapping is complex and it is not transparent how much information about the overall volume the different preprocessing procedures leave in the normalized data that could render the accuracies of different machine learning programs incomparable. Moreover, many studies do not describe such input normalization by overall brain volume and it must therefore be assumed that it has not been accounted for. In such a case the reported accuracy values have very limited relevance to the question we pose. For example, the recent studies of Xin et al. (2019) and Luo et al. (2019) have reported accuracies of 93% and 96.7% but have not described a correction for overall brain volume. Eliot et al. (2021) summarize in their review that 8 out of 12 studies on sex prediction did not correct for brain size. Three further studies reported a decline in accuracy to 60%-70% when brain size was controlled for with normalization (Eliot et al., 2021).

Two recent articles specifically studied the influence of a variety of TIV-adjustment methods when searching sex-specific regional differences (Sanchis-Segura et al., 2019) or when predicting sex using multivariate methods (Sanchis-Segura et al., 2020). The former study found that the TIV-adjustment method has a strong influence on the outcomes. In the later study, when no correction for TIV was performed, sex could be reliably predicted with >80% accuracy. However, after controlling TIV variation with the power-corrected proportions method, the prediction accuracy dropped to about 60%. Sanchis-Segura et al. (2020) even conclude that multivariate sex differences in gray matter volumes are largely dependent on male–female differences in TIV, a claim we dispute here using what Sanchis-Segura et al. (2019) refer to as the “only undisputed method to completely remove head-size variation”, a TIV-matched subsample.

Even a highly accurate sex classifying model or program is of limited relevance if it only performs well on the very same population of examples that it was trained on. Overfitting problems are usually accounted for by cross-validation or by partitioning example sets into training, validation and test sets. A little studied question is the robustness of a classifier, when the distribution of input data to be classified is different from the one the classifier was trained on. Such generalizability is important for any method that would eventually be applied clinically. Naturally, a possible loss in accuracy when predicting cross-cohort could vary widely with the dissimilarity of used scanners, in silico data preprocessing methods, and population differences, in particular with respect to age and provenience. Anderson et al. (2019) tested a weak form of generalizability using two cohorts. They studied a cohort of prisoners and a cohort of non-incarcerated people and obtained similar results on each. However, they trained in each case on the same data set they evaluated on and did not report any cross-cohort prediction accuracies. With regards to cross-cohort experiments, Eliot et al. (2021) found that “thus far, the only two [sex/gender] prediction studies to test their algorithms on external populations both found their accuracy to drop to near chance levels”. Joel et al. (2018) measured cross-data set performance on test cohorts from three geographical locations different from the one of the training set. The cross-cohort accuracies were between 71% and 86%. More recently, Sanchis-Segura et al. (2020) performed a cross-cohort (external) validation, when they trained on a subset of narrow age range from the HCP and used the trained models to predict on their own data set with similar age distribution. Thereby, logistic regression and a simple artificial neural network achieved accuracies of 62% and 57% only. The development of robust methods that perform well cross-cohort has been challenging.

In this work we demonstrate that sex can indeed be predicted with high accuracy from high resolution T1-weighted MR imaging, even when the effect of the total brain size is completely removed by matching males and females by TIV. We also introduce a certain 3D convolutional neural network, BraiNN, that is more accurate than logistic regression when the training cohort is smaller as well as more robust when predicting cross-cohort. In addition, we break down the importance of individual regions of interest (ROIs) for our multivariate classifier to test which brain areas are contributing most to the discriminatory power. BraiNN is open source and available from http://github.com/mabl3/BraiNN.

## 2. Methods

### 2.1. Characterization of data basis

In this study, we used two data sets from two different cohorts, the Human Connectome Project (HCP) and the Study of Health of Pomerania (SHIP). The SHIP data set includes data from the SHIP-2 and the SHIP-TREND-0 cohorts. Both cohorts include participants from the region of West Pomerania, Germany. SHIP-2 examinations were conducted from 2008 to 2012 and for SHIP-TREND-0 from 2008 to 2011 (Völzke et al., 2011). Both SHIP cohorts were pooled together, resulting in a data set of 3308 MRI scans. Participants ages ranged from 21 to 90 years with a mean of 53 years and an almost balanced sex (48.7% self-reported ‘male’ and 51.3% self-reported ‘female’). The overall mean TIV was 1545 ml (978 - 2311 ml), with 1646 ml for male and 1450 ml for female brains. The average TIV of men was 13.5% larger than that of women, matching approximately the aforementioned finding. The SHIP data set was primarily used for training and evaluation of the models.

To assess cross-population performance of the models, we used the HCP S1200 data set of the Human Connectome Project (Van Essen et al., 2013). The HCP data set contains 1113 T1w MRI scans, acquired from 2012 to 2015 on healthy young adults of age 22 – 37 (mean 29) and a near-balanced sex (with 45.5% self-reported males and 54.5% self-reported females). The overall mean TIV was 1441 ml (1017 – 1960 ml), with 1553 ml for male and 15.3% more than the 1347 ml for female brains. See Figure 1 for an illustration of data set properties.

**Figure 1:**
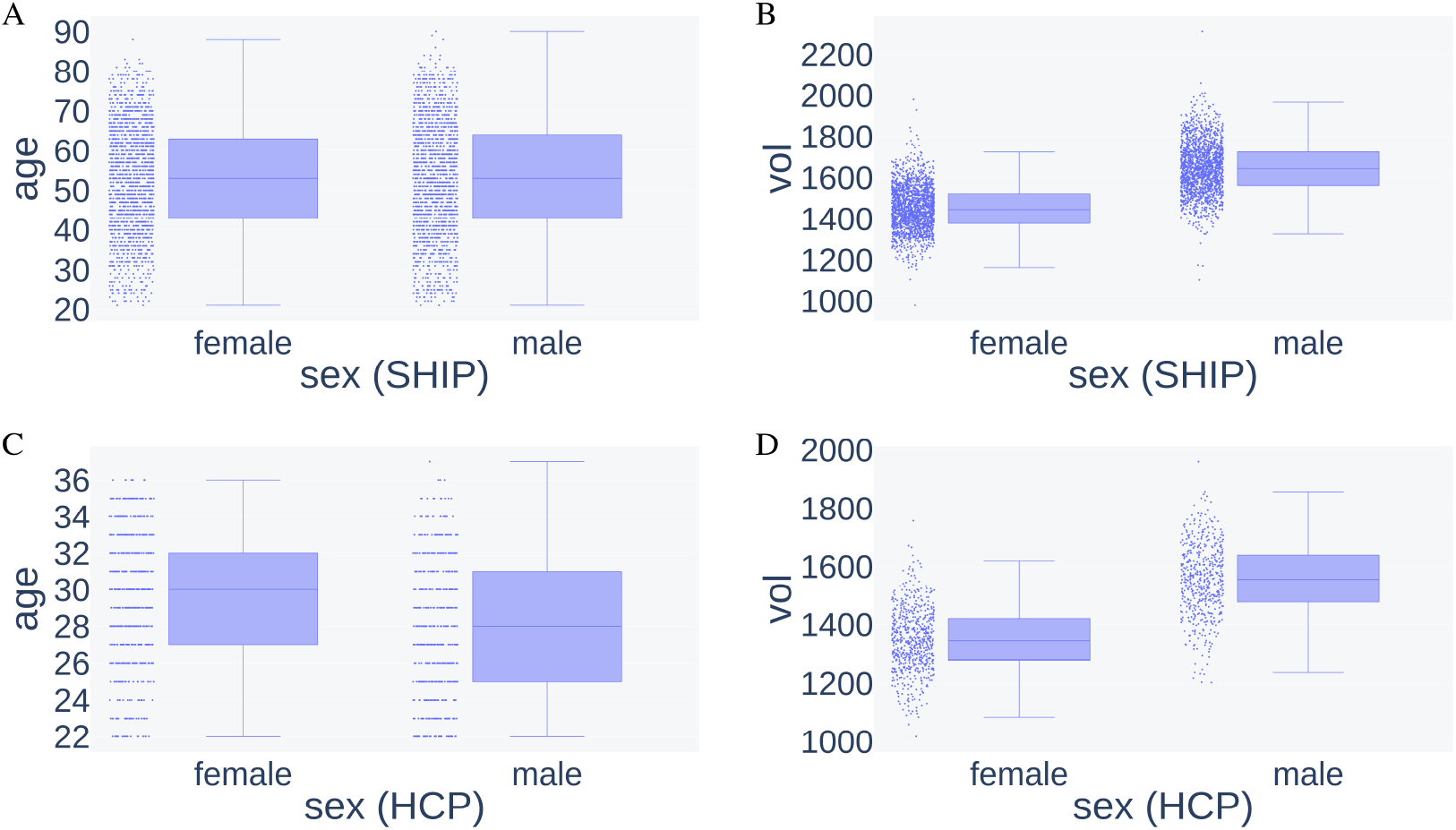
Data set properties age and TIV (vol) in male and female groups of both data sets. A) Age distribution in female and male samples in the SHIP data set. B) TIV distribution in female and male samples in the SHIP data set. C) Age distribution in female and male samples in the HCP data set. D) TIV distribution in female and male samples in the HCP data set.

The primary feature to be assessed by our proposed model was the voxel-wise gray matter volume (GMV) of high resolution T1-weighted anatomical MRI scans. For this purpose, tissue segmentation was performed using the SPM and CAT12 toolboxes running on the MATLAB platform. The following software versions were used: SHIP cohort data SPM v6225, CAT12 v1073; HCP cohort data SPM v7487, CAT12 v1450. At first, a spatial adaptive non-local means (SANLM) denoising filter (Manjón et al., 2010) improved signal-to-noise ratio, which was followed by internal resampling to properly accommodate low-resolution images and anisotropic spatial resolutions. The data were then bias-corrected and affine-registered to further improve the outcomes of the following steps, followed by the standard SPM “unified segmentation” (Ashburner and Friston, 2005). This acted as a starting point for the refinement stage. After skull-stripping, the brain was then parcellated into the left and right hemisphere, subcortical areas, and the cerebellum. Furthermore, local white matter hyperintensities were detected to be later accounted for during the spatial normalization. Subsequently, a local intensity transformation of all tissue classes was performed, which is particularly helpful to reduce the effects of higher gray matter intensities in the motor cortex, basal ganglia, or occipital lobe before the final adaptive maximum a posteriori (AMAP) segmentation. This final AMAP segmentation step (Rajapakse et al., 1997), which is independent of *a priori* information of the tissue probabilities, was then refined by applying a partial volume estimation (Tohka et al., 2004), which effectively estimates the fractional content for each tissue type per voxel, encoded as a probability value between 0 and 1 for each tissue type. As a last default step, the gray matter (GM) tissue segments were spatially normalized to a common reference space (MNI152 NLIN 6th generation) using DARTEL (Ashburner, 2007) registrations. For the HCP cohort prior to spatial normalization a cohort-specific template was calculated using the DARTEL template creation procedure of SPM. The spatially normalized GM images were modulated in a way, that each voxel encodes the local GMV prior to spatial normalization. As a spatial non-linear normalization removes most of the volume differences to the common template, the local volumetric changes due to this transformation have to be encoded. For this purpose the Jacobian determinant of the local transformations were used to modulate each voxel intensity, which afterwards encode dilation and contraction during that transformation to the common template space. As a result, after spatial normalization, the sum ∑_*j*_ *X*_*j*_ of all voxel values of an individual equals his or her total gray matter volume up to small errors in the scale of 1 cm^3^. Further, then every voxel coordinate of each participant’s GM tissue segment point to the equal anatomical location, enabling the statistical comparison of location and GMV.

Afterwards, a reference template based mask of GM tissue probability (thresholded to 0.5 and binarised) was applied to each single image to retain highly probable GM voxels only. Then, each single image was standardized with a Z-score normalization

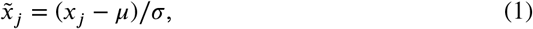

where *μ* and *σ* are the mean and standard deviation of voxel values in the image. Training and classification was performed on the final preprocessed images 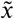.

Z-score normalization was important not only as a way of feature scaling for the machine learning algorithms, it also removed the correlation of the sum of voxel values in each image with the TIV. Before Z-score normalization, the voxel sum and TIV showed a positive correlation with Pearson’s correlation coefficient *r* = 0.645 (see Supplementary Figure 1). Through Z-score normalization, the voxel values in each image sum up to zero, which removes this correlation and is another important measure to rid our data from the nuisance variable TIV.

### 2.2. Machine learning classifiers

We compared two models for sex prediction, a convolutional neural network (CNN) and logistic regression. Both models are parametrized functions that take as input an image of dimension *m* = 121 × 145 × 121 and output a *‘femaleness’ score z* and a *‘femaleness’ probability p* = *σ* (*z*) ∈ [0, 1] (see Figure 5), where *σ* is the logistic sigmoid function, i.e. *p* = 1/(1 + exp(−*z*)). We classify a brain as female if and only if *p >* 0.5 or equivalently if the femaleness score is positive (*z >* 0) and otherwise as male. The *accuracy* is computed as the percentage of samples, where the predicted and actual sex agree.

The CNN, that is the basis of our BraiNN model, uses at its core the eponymous 3-dimensional convolutional layer. Further it uses pooling layers, a dropout layer for regularization and two fully-connected (‘dense’) neural network layers as depicted and detailed in Figure 2. The initial pooling layer was added to reduce the number of model parameters. We tried different pooling sizes and found that 6 × 6 × 6 pooling works well and with negligible impact on model performance (data not shown). The dropout rate in the dropout layer was 0.5. The activation function for the convolution and the 128 unit dense layer was PReLU (parametrized rectified linear unit). The second dense layer (the output layer) had a single unit with sigmoid activation function. In total, BraiNN has 1, 930, 625 parameters (or “weights”).

**Figure 2:**
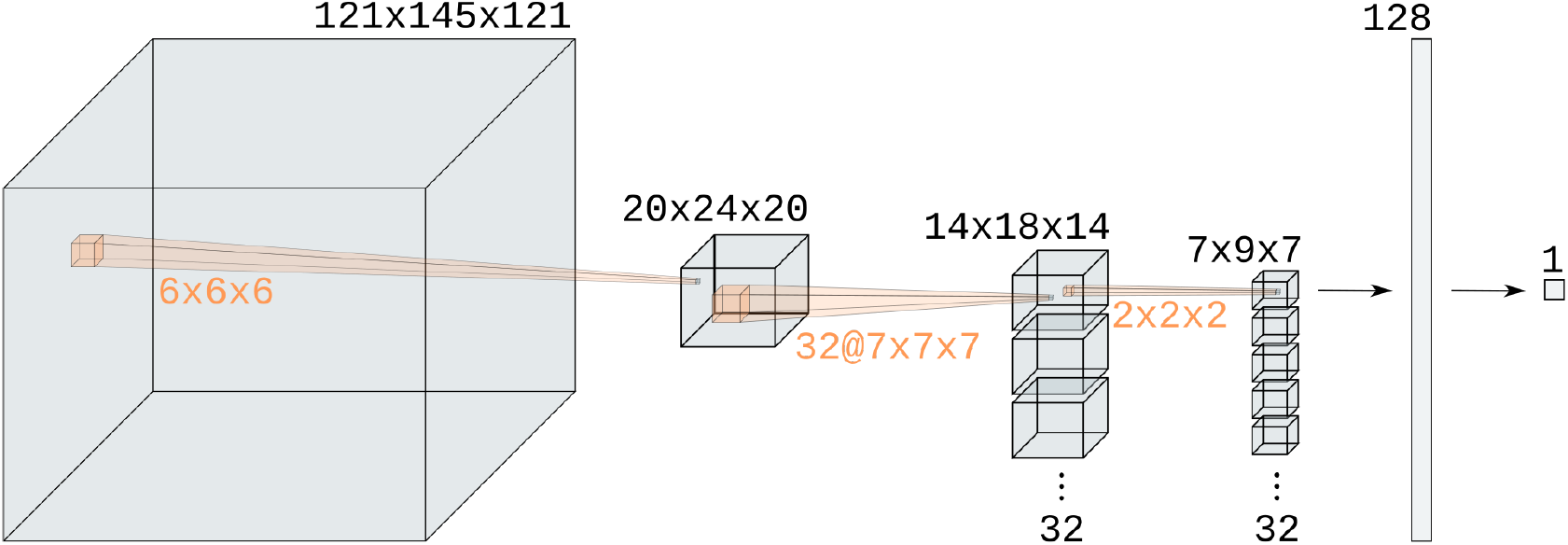
Illustration of BraiNN’s architecture. The layer dimensions are written above the layers, the pooling and filter dimensions in orange. The first layer is the input image, a 121 × 145 × 121 voxel MRI scan with one gray value per voxel. It is followed by a 6 × 6 × 6 max pooling (same stride), resulting in a 20 × 24 × 20 layer. To this, a 7 × 7 × 7 convolution with 32 filters is applied, yielding a 32 × 14 × 18 × 14 layer. This is again max-pooled with 2 × 2 × 2 (stride ‘same’), resulting in a 32 × 7 × 9 × 7 layer. This is flattened and fully connected to a 128 unit dense layer (left arrow). A dropout layer (not shown) with rate 0.5 is applied before the final dense layer (right arrow). The last layer outputs a single unit – the femaleness probability.

Logistic regression (here abbreviated as LogReg) uses weight *w*_*j*_ for each input voxel 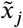 as well as a bias ameter *w*_0_ computes the femaleness score very simply as

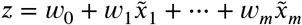

and therefore requires to train *m* + 1 = 2, 122, 946 parameters. As training criterion, the cross entropy error was minimized. The models were implemented using the TensorFlow framework (Abadi et al., 2015). Training used a stochastic gradient descent algorithm (Adam optimizer, learning rate 0.0001).

The BraiNN and LogReg parameters were estimated on the SHIP data set during a 5 × 5-fold cross-validation as described in the next section. BraiNN was trained for 100 epochs on the complete cohort data set and for 200 epochs on the matched data set, LogReg was trained for 30 epochs in both cases.

### 2.3. Cross validation

We used *k* × ℓ-fold cross-validation with *k* = ℓ = 5 for training, validation and evaluation. For this, the SHIP data set was randomly and under the uniform distribution split into five equally sized subsets. We then performed 5 “outer” training rounds, each time with a different one of the five subsets held back as a test set. The respectively remaining four subsets were combined to use for training. In each of the “outer” training rounds, we performed a nested cross validation by again randomly splitting the training data into 5 equally sized subsets. Again, five “inner” training rounds were performed, while holding back each time a different one of the five inner subsets as a *validation* set. The respectively four remaining inner subsets ((4/5)^2^ = 64% of the data) were combined to the actual training set. We then trained the model on the training set, using the validation set during training to monitor and control the training process. When the loss on the validation set did not decrease significantly (i.e. by more than 0.0001) for more than four epochs, the learning rate was reduced by multiplying it with 0.75. The model weights were stored each time the validation loss reached a new minimum. After training was finished, the model performance was evaluated on the test set. For this, the weights from the point of the lowest validation loss during training were used. At each training round, it is ensured that the test and validation data were not used for parameter estimation and that the test data was never seen during the training procedure. This way, we trained the model 25 times on different random splits of the data set, and report the average performance of the single models on their respective test data as result. Performing cross-validation reduces random effects where a single split of the data into training, validation and test set works particularly good or bad and gives a more confident estimate of how well the model will perform on upcoming new data.

### 2.4. Human expert classification

We challenged an experienced MRI-researcher and neurologist (author Martin Lotze) to compete against BraiNN in classifying the sex based on a subset of MRI scans. We randomly sampled 100 scans from the SHIP data set. He was allowed to look at the original complete MRI scans but sex information was withheld. We trained BraiNN on the remaining 3208 SHIP images and let it classify the same 100 images as Martin Lotze. BraiNN was only shown preprocessed images as before. The human expert principally had an advantage as the preprocessing for BraiNN removed helpful information such as TIV, head shape and the size of the ventricular system.

### 2.5. Correction for TIV sex differences through matching

It is well known that brain size, and for that matter body height, correlate with sex (Ruigrok et al., 2014). In fact, the self-reported sex in our SHIP data set can be classified with 80.7% accuracy when simply classifying any scan below a TIV threshold of 1551.91 ml as female and otherwise as male. Accuracies up to about 80% are therefore trivially achievable and irrelevant unless some correction for brain volume is performed.

Although the MRI scans are normalized for gray matter volume and should only encode local volume differences, we performed experiments on matched data sets to make sure the model does not benefit even from remains of global brain volume information. For this, the samples of the SHIP data set were grouped by sex and by TIV in steps of 10 ml. For each volume group, the larger of both sex subgroups was identified and a random sample of the same size as the smaller subgroup was taken. This way, we created a reduced ‘matched’ data set of 1274 images in which both sexes have virtually the same TIV distribution, shown in Figure 3, and cannot be distinguished based on that parameter any more.

**Figure 3:**
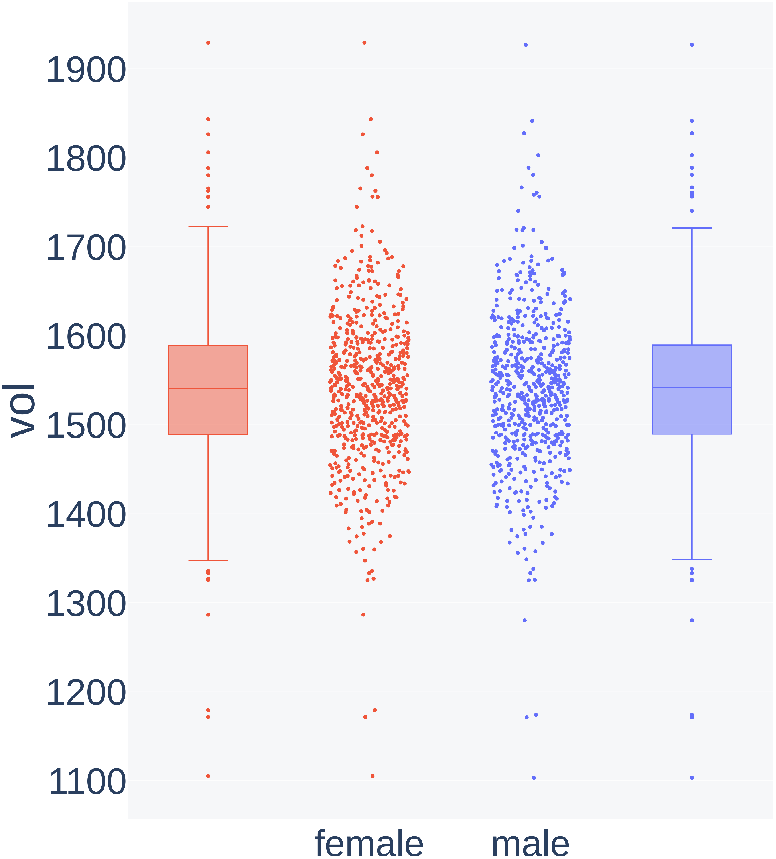
Distribution of TIV (vol, in cm^3^) in female and male MR images in the volume-matched data set.

Since the matched data set is much smaller than the complete cohort data set (39%), a reduced performance of a parameter rich model is to be expected simply from the fact that fewer training images were available. To assess this effect, we also created a reduced data set by randomly sampling 1274 images from the complete SHIP data set and trained our models on this reduced data set. To reduce stochastic effects, we repeated the random sub-sampling five times. Each time, training was performed with 5-fold cross validation. Here, the data is again randomly split into five equally sized subsets. Then, in five training rounds, each time a different subset is held back as test set and the remaining subsets are combined into the training set. This time, no validation sets are created for use during training. This way, the models were also trained 25 times on the reduced data set.

### 2.6. Regions of interest

To investigate the contributions of certain brain regions to the accuracy of sex prediction, we performed occlusion experiments respecting prior neuroanatomical knowledge. For this purpose we created 17 different binary regions of interest (ROI), compiled from 136 labels of the Neuromorphometrics atlas (provided by Neuromorphometrics, Inc. under academic subscription) as included in SPM12 and saved in the same dimensions as the MRI images. See Figure 7 for the list of regions and supplementary file `Neuromorphometrics_Derived_ROIs.xlsx` for an exhaustive overview. In one experimental setting (‘only ROI’), we applied each ROI mask to each input image in a way that only the respective brain region was left in the image, and the remaining voxels were set to zero. In a second setting (‘masked ROI’), we did the opposite, i.e. only setting the corresponding ROI voxels to zero and leaving the remaining image voxels unchanged. Thus, in the ‘only ROI’ setting, the classification results indicate how well the models perform on just small regions of the brain, and the ‘masked ROI’ classification results indicate, how much the models *rely* on a brain region. We use the area under the receiver operation characteristics curve (AUC) to evaluate the model performances. In the ‘only ROI’ results, a high AUC for one or more ROIs could indicate that the respective regions are sufficiently different between the sexes. In the ‘masked ROI’ results, a much worse performance compared to the models that used the whole brain images would mean that the respective region was key to successful classification and thus could also indicate that the remaining regions are of low information about the sex.

## 3. Results

### 3.1. Performance using complete cohorts

First we compared the two machine learning classifiers BraiNN and LogReg with each other using all – and therefore unmatched – data of the complete SHIP cohort as described in Section 2.3. Recall, that the spatial non-linear normalization described in Section 2.1 is supposed to control for overall brain size already. The average performances are shown in Table 1. LogReg performed better on the SHIP test data with 94.59% accuracy, but BraiNN also got a high accuracy of 89.78%. This is also mirrored in the area under the receiver operation characteristic curve (AUC). LogReg had an AUC of 0.988, BraiNN of 0.962 on the SHIP test data. Supplementary Figure 2 shows these ROC curves on the left.

**Table 1.** Mean accuracies of BraiNN and LogReg model in different training scenarios (’Training Data’). In the ’Complete SHIP’ case, the entire SHIP data set was used for training (see Section 2). For ’Matched SHIP’, the modified SHIP data set with equal TIV distributions for males and females was used (Section 2.5). In the ’Reduced SHIP’ scenario, randomly sampled subsets of the entire SHIP data set were used with the same size as the ’Matched SHIP’ data set. ’SHIP Accuracy’ denotes the fraction of correctly classified MRI scans from the respectively held-back test data. ’HCP Accuracy’ is cross-cohort and denotes the fraction of correctly classified MRI scans from the entire HCP data set after training on SHIP data only.

| Training Data | Model | SHIP Accuracy | HCP Accuracy |
| --- | --- | --- | --- |
| Complete SHIP | LogReg | 94.59% | 69.83% |
|  | BraiNN | 89.78% | 83.11% |
| Matched SHIP | LogReg | 90.91% | 71.45% |
|  | BraiNN | 86.15% | 73.34% |
| Reduced SHIP | LogReg | 92.87% | 65.26% |
|  | BraiNN | 88.40% | 80.60% |

The LogReg accuracy is just under the maximum reported accuracy we found in the literature: Feis et al. (2013) had reported a sex classification accuracy of 96%, achieved with a support vector machine. However, their study did not describe any control for overall brain size, indicating this comparison may not be fair. Further, their data set contained only *n* = 67 + 55 individuals, suggesting a relatively low precision of the accuracy estimate.

### 3.2. On volume-matched participants

In order to ensure that possible undetected remains of lobal brain volume information in the data do not distort the models, we then used the matched SHIP data set described in Section 2.5 for training of a LogReg and BraiNN. Both methods were evaluated using 5 × 5-fold cross-validation. As can be seen from the results in Table 1, LogReg still performed better on the SHIP test data with an accuracy of 90.91% compared to BraiNN with 86.15%. The AUCs for the matched data set show the same trend as with the complete cohort data set. LogReg has an AUC of 0.969, BraiNN of 0.930. See the left side of Figure 4 for the ROC curves.

**Figure 4:**
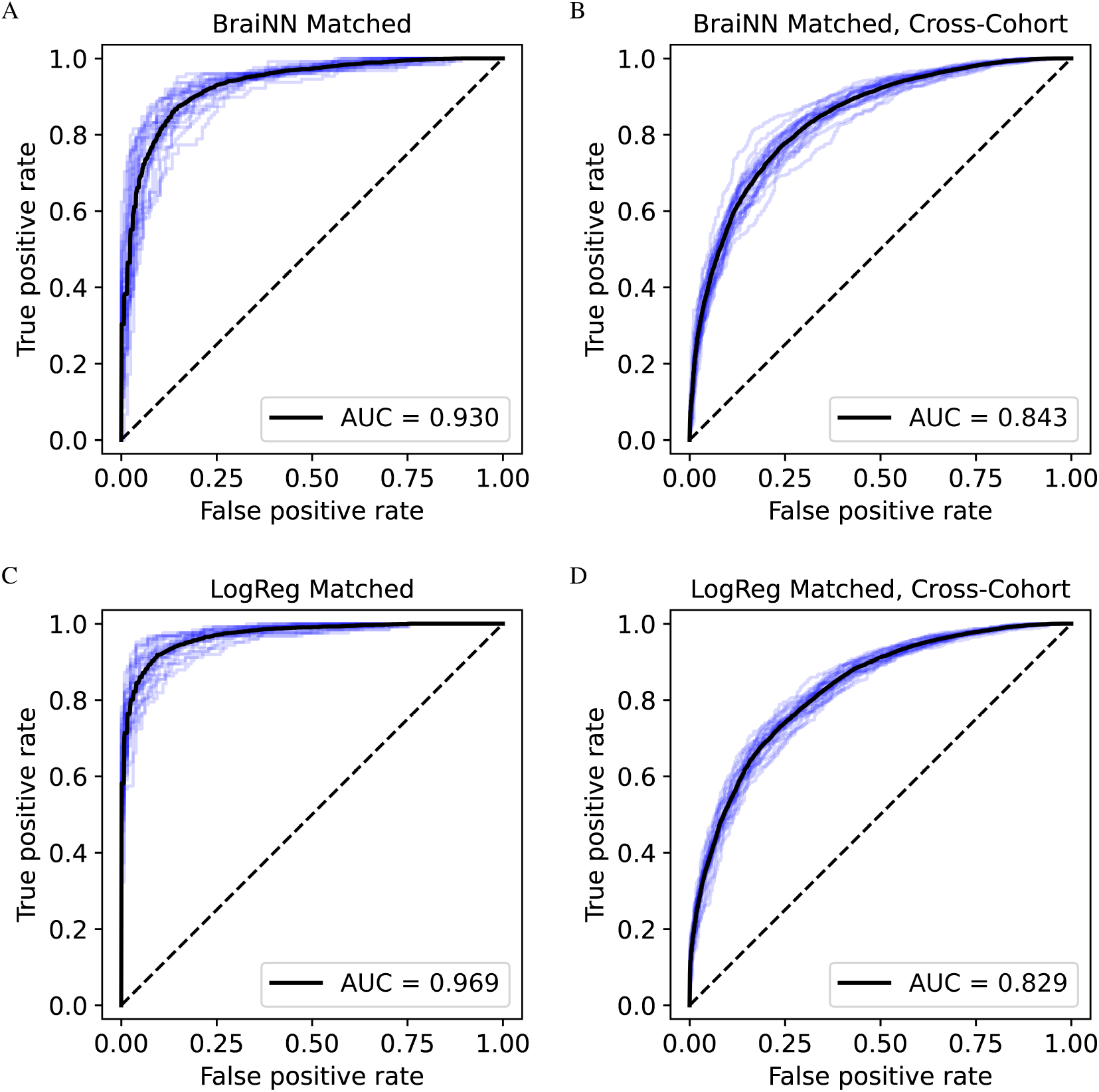
Receiver operation characteristic (ROC) curves for BraiNN (CNN) and LogReg on the matched SHIP data set. The ROC curves of each single training run are shown in blue, the black curves are the respective averaged ROC curves. The mean area under the curve (AUC) is shown in the bottom right of the plots. A) ROC and AUC for BraiNN, trained on the matched SHIP data set when predicting the SHIP test data. B) ROC and AUC for BraiNN, trained on the matched SHIP data set when predicting the HCP data set. C) ROC and AUC for LogReg, trained on the matched SHIP data set when predicting the SHIP test data. D) ROC and AUC for LogReg, trained on the matched SHIP data set when predicting the HCP data set.

The distribution of the femaleness score *z* and femaleness probability *p* output by BraiNN for the test images is shown in Figure 5. The femaleness score distribution for male images peaks at around -3 to -4 and for female images at around +3 to +4, both distributions appear to be approximately normal distributed. BraiNN is correctly and confidently predicting the sex of the majority of test images. There are few extreme cases of women classified as very likely male and vice versa. However, the shape of Figure 5A does not suggest that there is a substantial fraction of label errors, in which case either distribution would be expected to have a second mode. The femaleness probability is correctly close to 0 or 1 for most images and the default classifier threshold of 0.5 appears to be a reasonable choice in this setting.

Previously, Wang et al. (2012) had performed sex classification on a volume-matched data set of only 35 male and 35 female subjects. Their support vector machine had as input combined structural MRI and resting-state fMRI data and obtained a mean classification accuracy of 89%. Our similar accuracy confirms the result that even with a careful gold-standard control for brain volume the sexes can be distinguished quite accurately. Furthermore, our much larger data set of 1274 images allows for more precise estimates of the accuracy and we here used only part of the input features that Wang et al. (2012) had used.

### 3.3. Influence of training cohort size

Using a volume-matched subset in Section 3.2 reduced the prediction accuracy of both studied methods with respect to using the complete SHIP cohort in Section 3.1. As hypothesized, this could be a consequence of what is sometimes called *feature leakage* in the machine learning domain. This could be the case if at least partial information from the total brain volume is still represented in the complete cohort data set, even though some normalization method was applied. On the other hand, smaller training sets also generally lead to less accurate models, when their number of parameters is large. To assess the impact of the latter, i.e. of the reduced number of training images, we also trained the models on randomly sampled subsets from the SHIP data set. As one of the subset sizes we chose the size of the matched data set, 1274, in order to make comparisons that allow to disentangle the two expected causes for said difference.

Figure 6 shows the accuracies of our methods as a function of cohort size. Note that with the cross-validation we use, a larger cohort size both allows a better parameter training and a more precise evaluation of the accuracy.

**Figure 5:**
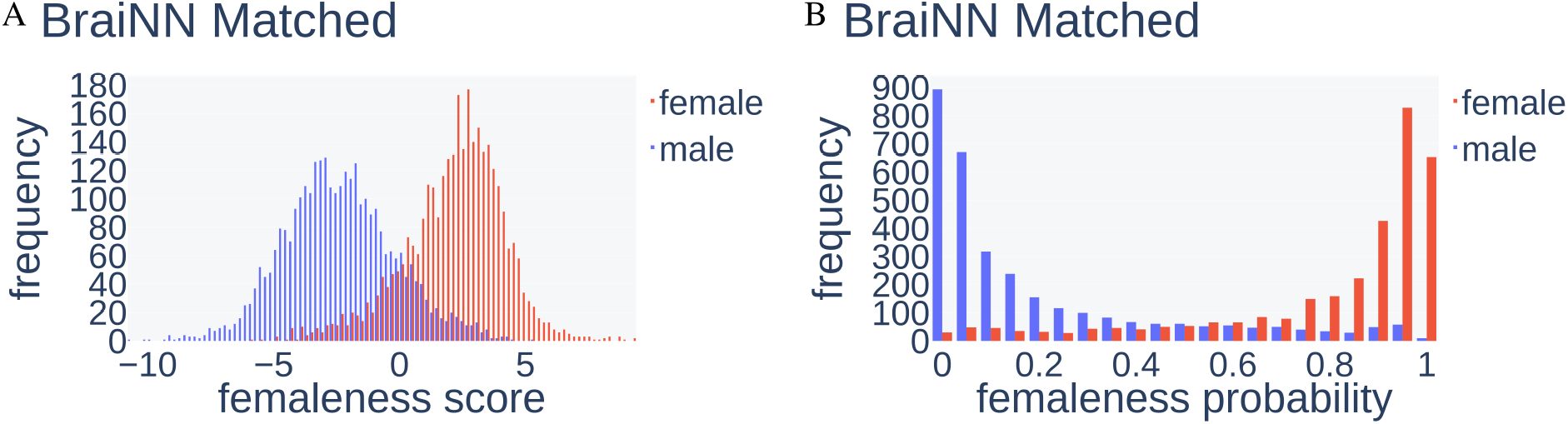
Distribution of femaleness score and femaleness probability from BraiNN on SHIP test images after training on the matched SHIP data set. A) Distribution of the femaleness score, red bars depict the frequency (y axis) of the score (x axis) for female scans, blue for male scans. A score below 0 leads to classification of the image as male, otherwise as female. B) Distribution of the femaleness probability. Scans with a femaleness probability above 0.5 are classified as female.

**Figure 6:**
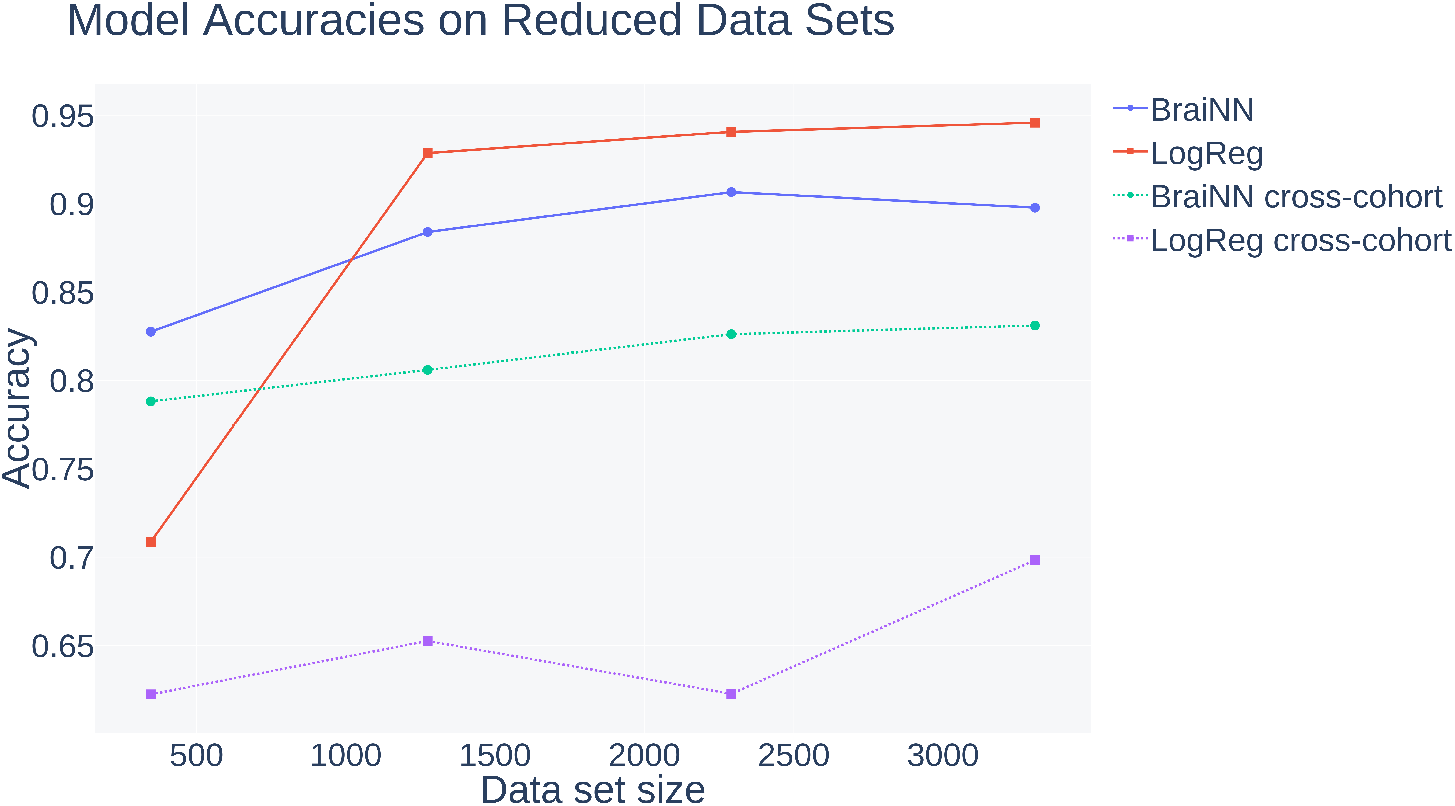
Influence of the data set size (347, 1274, 2291 and 3308) on the model accuracies. The smaller sized data sets were randomly sampled from the complete cohort data set and training was performed as described in Section 2.5.

**Figure 7:**
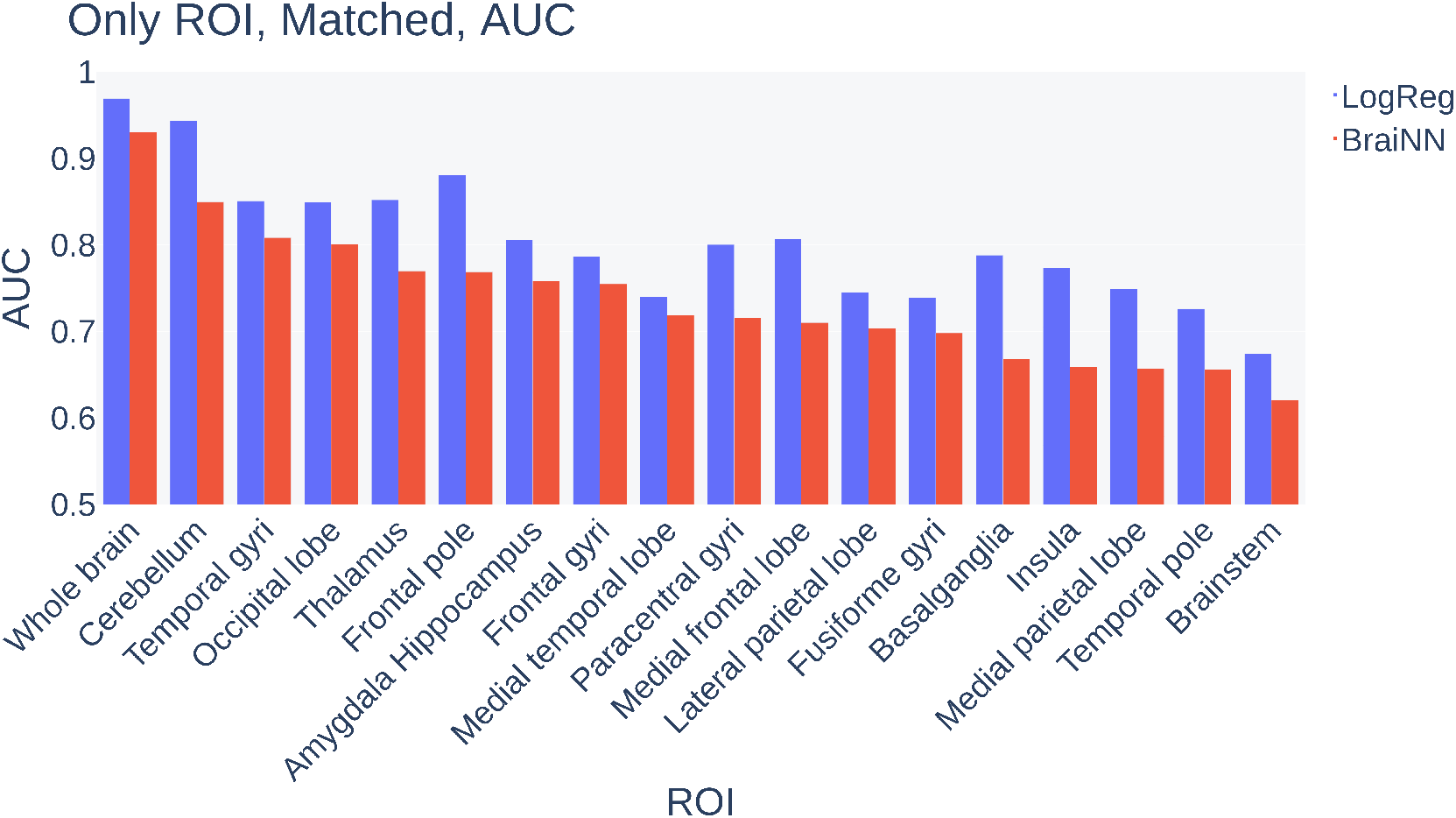
AUC of LogReg and BraiNN for SHIP images of “whole brain” or only a certain ROI. The volume-matched SHIP data set was used for training.

In comparison, the performance on the randomly reduced data set of the same size of the matched data set was 92.87% accuracy for LogReg and 88.40% for BraiNN on the SHIP test data. Thus, less than half of the lost percent points from the whole data set training to the matched data set training can be explained by the reduced training set size. If we subtract the losses due to a smaller (random subset) cohort size from the losses due to using a volume-matched data set, we obtain estimates of the effect of feature leakage: For BraiNN, an estimated (89.78−86.15)−(89.78− 88.4) = 2.25 percent points and for LogReg an estimated (94.59 − 90.91) − (94.59 − 92.87) = 1.96 percent points of the loss in accuracy when using a volume-matched data set rather than a normalization approach can be attributed to an incomplete accounting for TIV. This confirms quantitatively that matching for TIV is the gold standard for removing the influence of body size on brain comparisons between males and females. Naturally, if not done with matching, the choice of a brain-size normalization procedure may have a large influence on sex predictability (Sanchis-Segura et al., 2020).

The leftmost data points in Figure 6 show the accuracy for a data set size of 347, which is the number of children of which Kurth et al. (2021) classified their sex using relevance vector regression (RVR) following a principal component analysis (PCA). On their data set Kurth et al. achieved a classification accuracy of 80.4%. When we downsample our data set to the same size in order to correct for advantages from our larger data set, BraiNN achieves an accuracy of 82.8%. This suggests that our convolutional neural network is competitive with or better than the approach of Kurth et al. (2021).

The solid curves crossing each other in Figure 6 show also very prominently that BraiNN is more accurate than LogReg on smaller data sets below roughly 1000 participants. Logistic regression, as performed by us, looses accuracy very quickly when the cohort size is small and – although better for large cohorts – would likely be noncompetitive for many of the cohort sizes from the relevant literature.

### 3.4. Cross cohort prediction

A major concern for translation of machine learning (ML) methods to clinical use is their robustness with respect to differences in machines, protocols, preprocessing software and practices. A ML method could perform better than a human expert when applied to data from the same distribution as the data it was trained on but perform poorly and much worse than an expert when applied to data, say, from a different machine. In order to study the robustness of our methods we used a second data set with data from the Human Connectome Project (HCP), which is unrelated to the SHIP cohort and its age distribution is narrower and with a much smaller mean. This allows to assess how well the trained models generalize to new images from a different scanner and population. We then performed within-data set and between-data set experiments. In the first, SHIP images were used both for training and performance evaluation, in the later, SHIP images were used for training and HCP data for performance evaluation. In both cases 5 × 5-fold cross-validation was performed. Table 1 shows the results for logistic regression and our CNN.

As expected, in a cross-cohort prediction the accuracy drops for both methods compared to the within-cohort results (Table 1). When training on the complete SHIP cohort and predicting on HCP, the LogReg performance steeply dropped to 69.83% accuracy (AUC 0.919, see right side of Supplementary Figure 2). In the cross-cohort predictions, BraiNN retained a higher accuracy of 83.11% (AUC 0.927). When trained on the volume-matched SHIP data set, the accuracies on the HCP cohort were 71.45% for LogReg and 73.34% for BraiNN (AUC 0.829 and 0.843, respectively, see right side of Figure 4).

The training set size had a smaller impact when training cross-cohort, presumably as the performance is overall lower and a good fit to the training cohort is not necessarily an advantage (Figure 6). BraiNN is consistently more accurate than LogReg by about 15 percent points, e.g. the HCP performance dropped to 65.26% for LogReg and 80.60% for BraiNN when trained on a random subset of size 1274 of the SHIP data set. Overall these experiments in which BraiNN always performed more accurately than LogReg suggest that our BraiNN convolutional neural network is more robust than logistic regression towards the changes in the data set distribution stemming from changes in the scanner, its software and the population.

### 3.5. Regions of interest

We performed the ROI experiments on both the complete cohort and matched SHIP data set. We observed overall the same effects in both cases and will therefore only describe the matched data set results in more detail. The other results can be found in the Supplementary Material. Note that in some experiments we found a poor model accuracy but a rather high AUC. For example, using only the cerebellum for training resulted in an accuracy of 0.483 for LogReg when using the default classifier threshold of 0.5 (see Section 2.2), while the AUC was 0.824. Here we observed a tendency to predict values close to zero for both males and females (not shown). With a classifier threshold of 0.5, this then leads to almost every image being predicted as male and thus the poor accuracy. However, the high AUC indicates that LogReg was indeed able to accurately predict the test data with a better suited classifier threshold. We thus here compare the ROIs based on the more meaningful AUC measure rather than the accuracies.

The first experimental setting ‘only ROI’, where only one region is ’visible’ to the classifier, revealed that for both models any of the investigated single brain regions are enough to classify brains with an AUC of at least 0.620. While slightly better than random guessing, this is a poor performance compared to the whole brain images, where LogReg and BraiNN achieve 0.969 and 0.930, respectively. However, some single ROIs performed quite well on their own. Especially the cerebellum showed the highest AUC for both models with 0.943 and 0.849 for LogReg and BraiNN, respectively. When classifying the HCP data set, the cerebellum also performed best with an AUC of 0.824 and 0.759 for LogReg and BraiNN (whole brain images: 0.829 (LogReg) and 0.843 (BraiNN)). See Figure 7 for a bar plot comparing the AUC of the “whole brain” images and the single ROIs for both models. Supplementary Figure 3 shows a bar plot of the model performances on the HCP data set. The exact values are shown in Supplementary Table 2. Other regions with an AUC *>* 0.8 (LogReg, BraiNN) were temporal gyri (0.850, 0.808) and occipital lobe (0.849, 0.801).

The second experimental setting (‘masked ROI’) showed for both models and both training data sets only slight differences in model AUC, i.e. leaving out any ROI could be compensated by the models and performed almost as good as when the whole brain was input. See the Supplementary Material for bar plots illustrating the different AUCs for “whole brain” images and images with masked ROIs for both models and both training data sets, and the performances on the cross cohort data (Suplementary Figures 2, 4 and 6). The exact values are shown in Supplementary Tables 3 and 4.

### 3.6. Machine versus human expert

Of the 100 test scans given both to BraiNN and Martin Lotze, BraiNN correctly classified 87 scans and ML correctly classified 62 scans. 36 images were correctly classified by the program and not by the expert, whereas 11 images were correctly classified by the human and not by the program, 51 were correctly predicted by both. BraiNN is more accurate at sex classification than this expert (*p <* 0.0004, two-sided sign test).

## 4. Discussion

We examined the question whether two same-size brains, one of a male and one of a female, are distinguishable from magnetic resonance images and how the sex classification accuracy of a machine learning model depends on the method, the cohort size and whether the method’s parameters were estimated on the same or a different cohort. The extent to which the brains of men and women differ had been studied extensively and discussed controversially. Recently, it has been argued that the human brain is not sexually dimorphic (Eliot et al., 2021), that the sex differences can largely be attributed to male-female differences in TIV and that TIV-adjustment methods can be ineffective and influence much the results (Sanchis-Segura et al., 2020). Analyses in the MRI domain of machine learning applications are also impeded by the fact that one team of researchers does (can) not reproduce the results of another team of researchers. Comparisons become inconclusive, e.g. when another cohort is used, other scanners, other TIV-adjustment methods as well as other classification methods and if the respective program’s code, version and calling parameters are not available.

We found that indeed same-size brains of men and women can be distinguished with high accuracy. At least an accuracy of about 91% can be achieved on a data set where males and females of the same TIV were matched. Thereby, the differences between the sexes cannot be explained away by differences in overall volume. In our large data set of 1274 volume-matched participants this accuracy also cannot be explained by chance. Therefore, the information to distinguish the brains of men and women is contained somehow in their MR images. Our methods achieve a better accuracy than a human expert but have the disadvantage that they do not intrinsically explain their decision.

We tried to order the relative importance of different regions of the brain for sex discrimination with our artificial neural network BraiNN by occluding ROIs as input to BraiNN during prediction and also during *training*. We found that cerebellum, temporal gyri and occipital lobe are contributing most to the discriminatory power. Interestingly, these have been the areas exhibiting larger GMV in males than in females in a previous paper (Table 2, Lotze et al. (2019)) and have been connected to sensorimotor (cerebellar hemispheres), (higher) visual recognition and perception (fusiform gyrus and occipital lobe) but also to cognitive processing (STS, temporal pole). However, the differences between brains of females and males are likely not restricted to size differences of certain regions of interest but more complex. None of the regions are by themselves necessary for discrimination and nearly the same accuracy can be achieved when each region is occluded versus when the whole brain is visible. We believe that more advanced methods for *explainable* artificial intelligence are required to assist an expert in interpreting sex differences beyond mere regional size differences, to explain why a particular brain belongs to the predicted class and to fill the gap of unexplained differences that statistical methods of perregion GMV leave.

**Table 2.** ROI based accuracy (AUC) of correct sex recognition from the BraiNN model, trained and tested on the volume-matched SHIP data set. Without any restricting to ROIs (’whole brain’ at bottom) BraiNN achieved an AUC of 0.93. When restricting BraiNN for instance on the cerebellar ROI an AUC of about 0.85 was reached. Interestingly, those areas with best differentiation performance were remarkably congruent to those which showed highest sex-differences in conventional VBM-comparisons (see Cohen’s d in last column averaged over both hemispheres and selected from both comparisons (GMV: “women>men” and “men> women”) from (Lotze et al., 2019).

| ROI | BraiNN<br>SHIP<br>AUC | Cohen’s D |  |
| --- | --- | --- | --- |
| Cerebellum/<br>Anterior cerebellar hem. | 0.85 | 0.33 | m>w |
| Temporal gyri/<br>Superior temporal sulcus | 0.81 | 0.27 | w>m |
| Occipital lobe | 0.80 | 0.29 | m>w |
| Thalamus | 0.77 | 0.21 | w>m |
| Frontal pole | 0.77 | 0.34 | w>m |
| Amygdala Hippocampus | 0.76 | 0.43 | m>w |
| Frontal gyri/<br>dlPFC: BA45 | 0.75 | 0.32 | w>m |
| Medial temporal lobe/<br>Parahippocampal gyrus | 0.72 | 0.53 | m>w |
| Paracentral gyri | 0.72 | 0.27 | m>w |
| Medial frontal lobe | 0.71 | 0.32 | w>m |
| Lateral parietal lobe | 0.70 | 0.25 | w>m |
| Fusiforme gyri | 0.70 | 0.36 | m>w |
| Basalganglia/<br>Putamen | 0.67 | 0.32 | m>w |
| Insula/<br>posterior insula | 0.66 | 0.28 | w>m |
| Medial parietal lobe | 0.66 | - |  |
| Temporal pole | 0.66 | 0.33 | m>w |
| Brainstem | 0.62 | - |  |
| Whole brain | 0.93 |  |  |

We show how the classification accuracy increases with the size of the data set available for training. In particular for smaller cohorts, say of 1000 participants or below, much accuracy is lost. This means that some of the true sex differences may not be uncoverable in studies on smaller cohorts. This also means that comparisons between methods should take different data set sizes into account. Correcting for the accuracy lost due a training set size reduction when going from a complete cohort to a volume-matched subset, we find that indeed – and despite our best effort – our complete cohort data set appeared to suffer from feature leakage, where our preprocessing with spacial and Z-Score normalization did not succeed to fully remove the information from the nuisance variable TIV.

An ultimate goal may be that a machine learning method can produce valuable information from an MRI scan, without requiring that thousands of other individuals from the same population have been scanned before on the same machine. Toward that goal we studied the loss in accuracy that is obtained when training on one cohort and evaluating on another cohort. Interestingly, the method that is more accurate on the same cohort (logistic regression) is less accurate when applied cross-cohort. The BraiNN convolutional neural network appears to be more robust with respect to the change of scanner and population, even though both methods have roughly the same number of parameters, about two million. After training on the complete SHIP cohort, BraiNN achieves an accuracy of 83.1% when predicting on the HCP cohort compared to 89.8% (cross-validated) when predicting on SHIP itself. Although this indicates that a convolutional neural network may generalize better than logistic regression, such results are likely sensitive to distributional differences between the cohorts and the generalizability would ideally be tested using scans from many or heterogeneous sources.

Previously, machine learning methods have been compared against humans for the medical image classification task. Schaffter et al. (2020) reported results on the Digital Mammography DREAM challenge where mammogram images were to be classified: Will the woman be diagnosed with breast cancer in the 12 months after taking the image or not? The competing automatic methods from 2017 were still substantially less accurate than a particular radiologist. In the broader ImageNet challenge, however, where photos from everyday life were to be classified into one of 1000 categories, the best machine learning method achieved near-human performance in 2015 (Russakovsky et al., 2015): The GoogLeNet convolutional neural network achieved a top-5 error of 6.8%, better than one human annotator, but yet worse than another (5.1%). The top-5 error rate is the percentage of images where the true class is among five predicted classes. Since, automatic methods have improved steadily and significantly and automatic methods have lowered the top-5 error rate for the ImageNet data to about 1.2% (Pham et al., 2021). It can be expected that ongoing improvements to ML methods of computer vision will translate to higher accuracies in classifying MR images as well.

### Limitations of the study

The ROIs we chose for the occlusion experiments are rather coarse. Further examinations could possibly improve interpretability by using a finer partition of the brain or a ’searchlight’ approach (Weaverdyck et al., 2020). Another limitation is that the sex attribute was self-reported and not complemented by sex-specific measurements, such as those of testosterone or oestrogen. We carefully excluded the information that brain size holds on sex. Also, male and female individuals had very similar age distributions in the SHIP cohort. Furthermore, the broad differentiation of ROI-specific recognition rates are an argument that neuroanatomy is indeed the factor modulating correct classification. However, another limitation is that we cannot exclude the possibility that some other further attribute of individuals – unknown to us or unrecognized – correlates with sex, is detectable in MR images but is irrelevant to the question how male and female brains differ principally.

## 5. Conclusions

The current study set out to investigate if sex can be predicted from gray matter volumes when total brain size is completely removed by matching males and females by total intracranial volume. In a large data set, we were able to achieve high classification rates also when evaluated on another cohort. Machine learning is able to find differences between brains of males and females that may not appear when using statistical models. Our convolutional neural network BraiNN achieves a comparably good accuracy already with a few hundred training images.

## Supporting information

regions of interest

## Data Availability

The source code of BraiNN is available from GitHub.

https://github.com/mabl3/BraiNN

## 6. Contributions

ME and MS planned the study, performed the analyses and wrote the manuscript. MD and ML planned the study and wrote the manuscript. NN performed the literature search and wrote the manuscript.

## 7. Funding

This project had been supported by a starter grand from „Nordeutsche Universitäten” and funds for digital education from the German Pact for Higher Education.

## Notes

### Competing Interest Statement

The authors have declared no competing interest.

### Funding Statement

This project had been supported by a starter grand from "Nordeutsche Universitaeten" and funds for digital education from the German Pact for Higher Education.

### Author Declarations

Ethics Committee of the University Medicine Greifswald gave ethical approval for this work

