## Supplementary Tables and Figures for "Classifying sex with MRI"

### Classifying sex with MRI
### Supplementary Material

Matthis Ebel[1], Martin Lotze[2], Martin Domin[2], Nicola Neumann[2], and Mario Stanke[1]

[1]Institute for Mathematics and Computer Science, University of Greifswald, Walther-Rathenau-Str. 47, 17489, Greifswald, Germany
[2]Institute of Diagnostic Radiology and Neuroradiology, Functional Imaging, University Medicine Greifswald, 17489, Greifswald, Germany

April 27, 2022

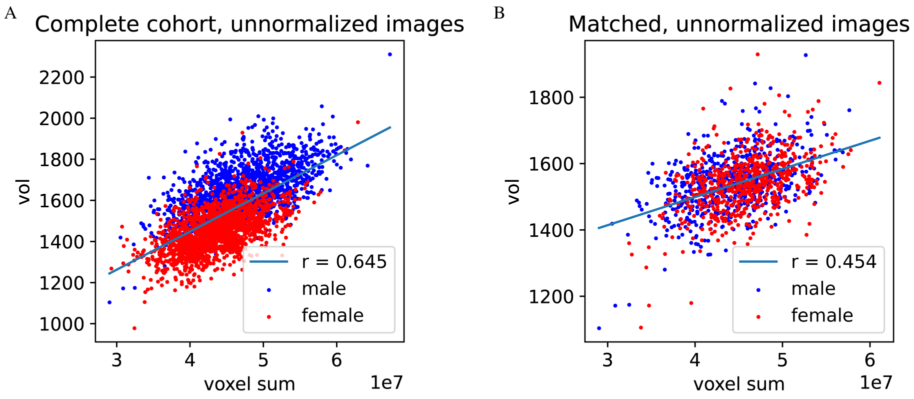


**Supplementary Figure 1:** Correlation of voxel sum in each MR image with the TIV (vol). Blue dots represent male samples, red dots female. The straight line shows a linear regression calculated on all images (male and female pooled), 'r' is Pearson's correlation coefficient. A) Correlation for the complete SHIP data set for images that have not been Z-score normalized. B) Correlation for the TIV-matched data set for images that have not been Z-score normalized.

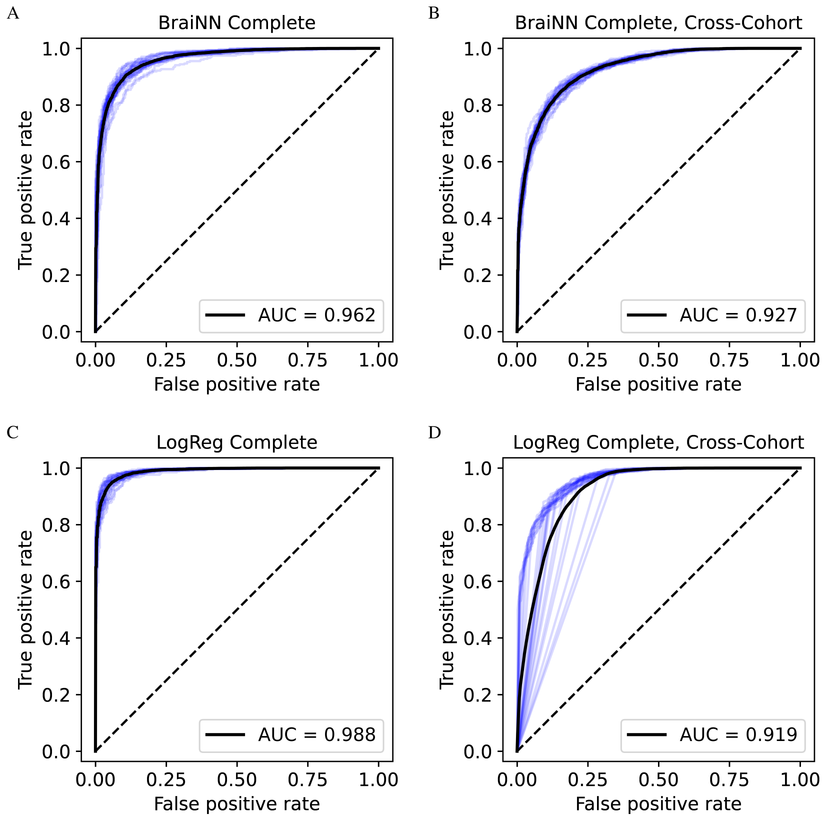


**Supplementary Figure 2:** Receiver operation characteristic (ROC) curves for BraiNN (CNN) and LogReg on the complete SHIP data set. The ROC curves of each single training run are shown in blue, the black curves are the respective averaged ROC curves. The mean area under the curve (AUC) is shown in the bottom right of the plots. A) ROC and AUC for BraiNN, trained on the complete SHIP data set when predicting the SHIP test data. B) ROC and AUC for BraiNN, trained on the complete SHIP data set when predicting the HCP data set. C) ROC and AUC for LogReg, trained on the complete SHIP data set when predicting the SHIP test data. D) ROC and AUC for LogReg, trained on the complete SHIP data set when predicting the HCP data set. The LogReg in this setting has a tendency to predict values close to 1, even for male images (data not shown), thus the shape of the blue curves here.

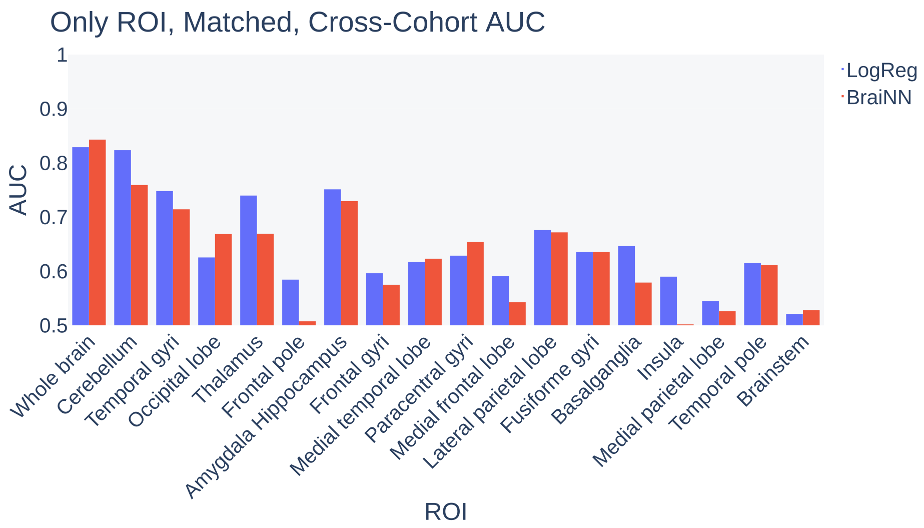


**Supplementary Figure 3:** AUC of LogReg and BraiNN (CNN) for "whole brain" images and images *missing* certain brain regions when predicting the HCP data set. In all cases, the matched SHIP data set was used for training. See Table 2 for the exact values.

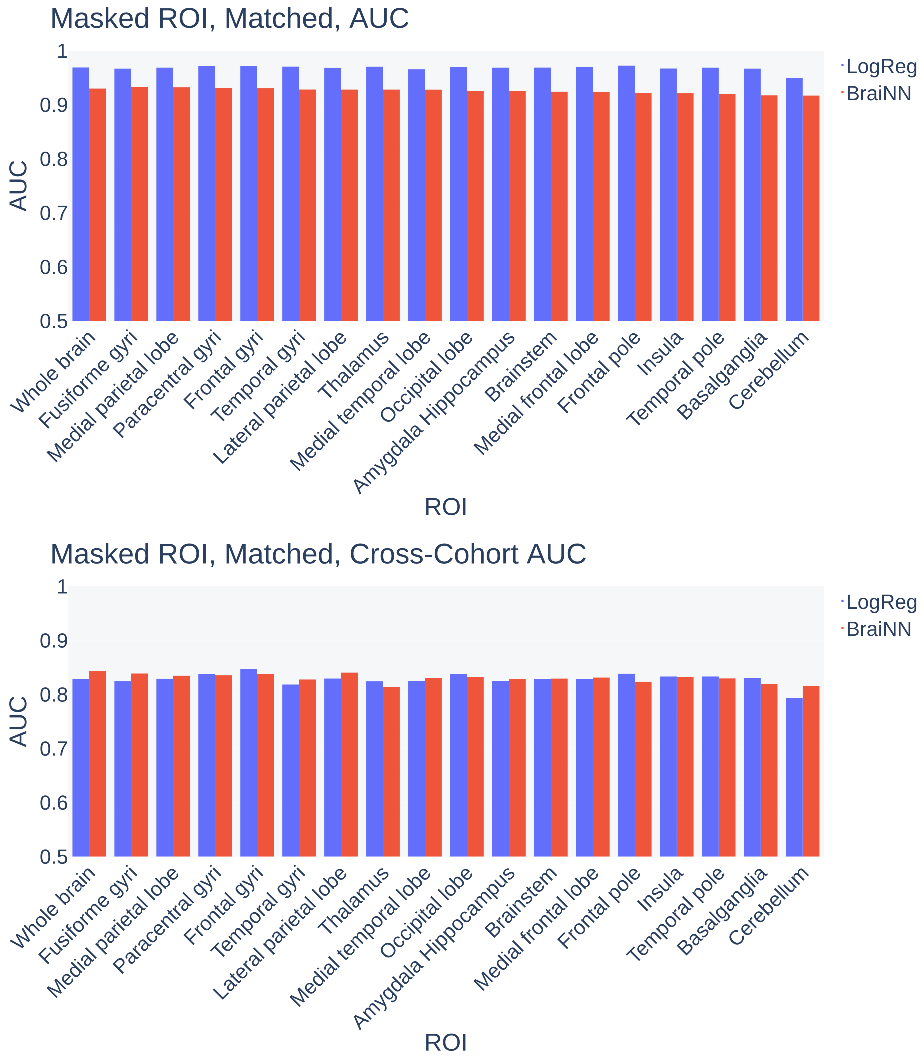


**Supplementary Figure 4:** AUC of LogReg and BraiNN (CNN) for "whole brain" images and images *missing* certain brain regions. Top: Performance on the test data (see Section 2.3). Bottom: Performance on the HCP data set. In all cases, the matched SHIP data set was used for training. See Table 4 for the exact values.

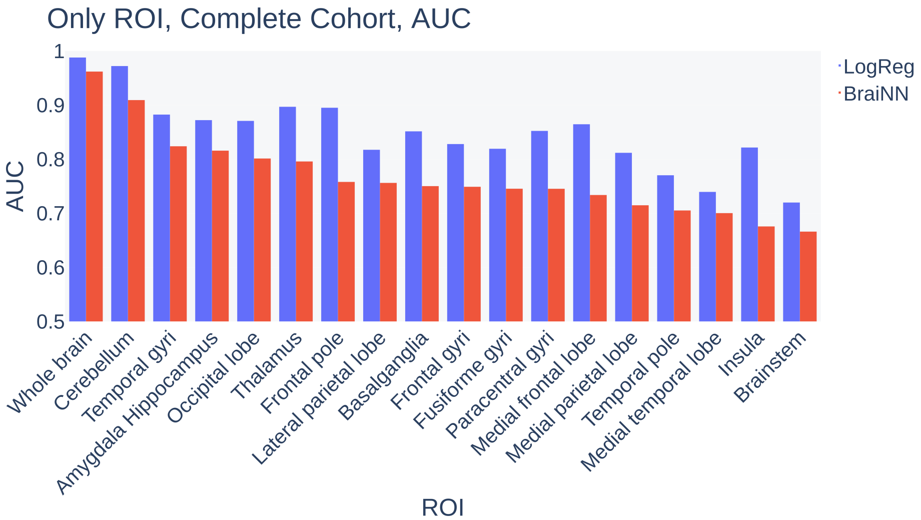


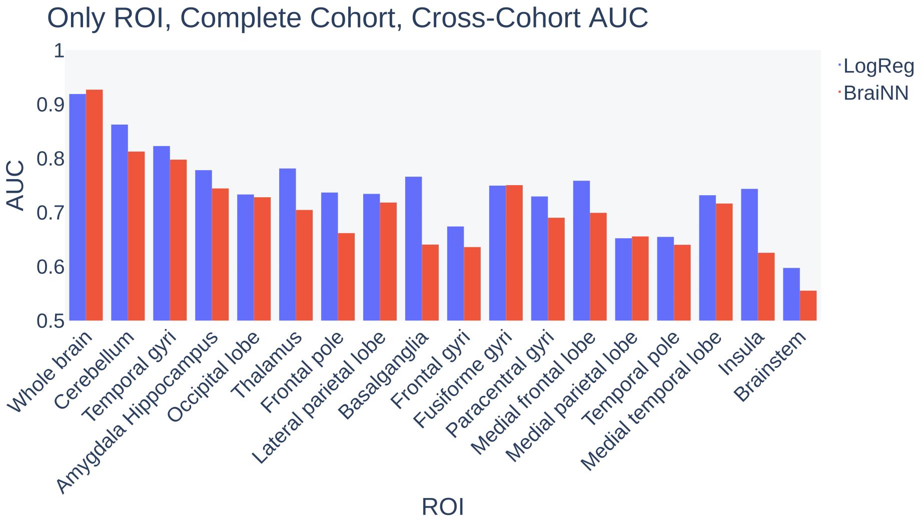


**Supplementary Figure 5:** AUC of LogReg and BraiNN (CNN) for “whole brain” images and images containing only certain brain regions. Top: Performance on the test data (see Section 2.3). Bottom: Performance on the HCP data set. In all cases, the complete SHIP data set was used for training. See Table 1 for the exact values.

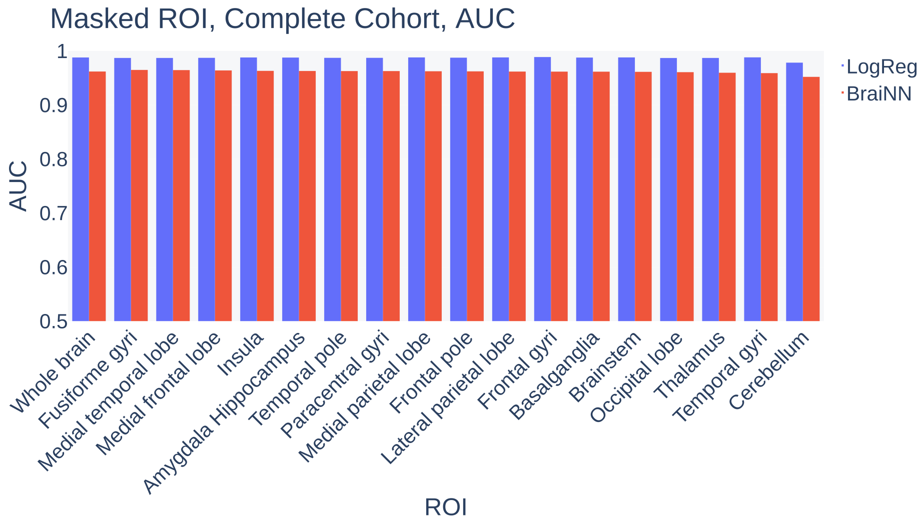


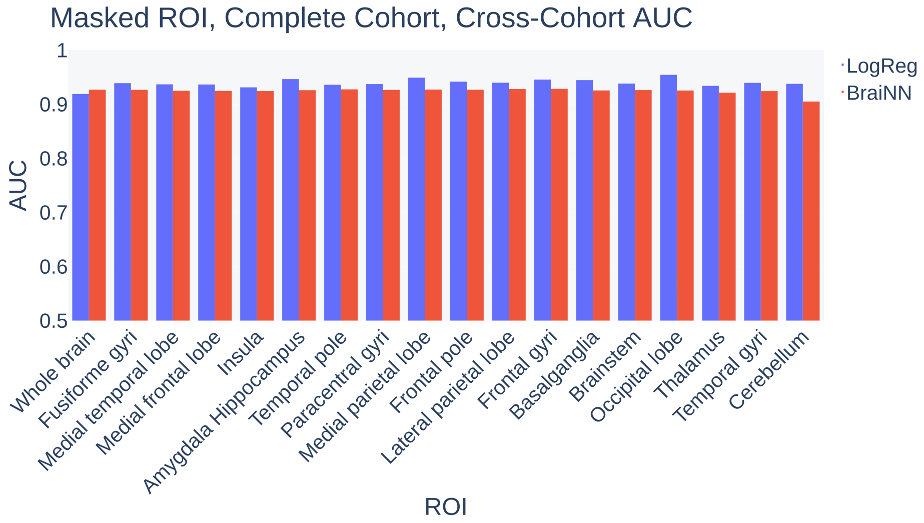


**Supplementary Figure 6:** AUC of LogReg and BraiNN (CNN) for "whole brain" images and images *missing* certain brain regions. Top: Performance on the test data (see Section 2.3). Bottom: Performance on the HCP data set. In all cases, the complete SHIP data set was used for training. See Table 3 for the exact values.

**Supplementary Table 1:** Accuracies and AUC for both models trained on the **complete SHIP** data set when only **certain ROIs are visible** in the images.

| **ROI** | **LogReg** | | | | **BraiNN** | | | |
|---|---|---|---|---|---|---|---|---|
| | **SHIP** | | **HCP** | | **SHIP** | | **HCP** | |
| | **accuracy** | **AUC** | **accuracy** | **AUC** | **accuracy** | **AUC ↓** | **accuracy** | **AUC** |
| Cerebellum | 0.9108 | 0.9722 | 0.6674 | 0.8625 | 0.8285 | 0.9093 | 0.7151 | 0.8126 |
| Temporal gyri | 0.8008 | 0.8825 | 0.7271 | 0.8228 | 0.7468 | 0.8239 | 0.6962 | 0.7976 |
| Amygdala Hippocampus | 0.7911 | 0.8722 | 0.6480 | 0.7782 | 0.7374 | 0.8158 | 0.6430 | 0.7444 |
| Occipital lobe | 0.7857 | 0.8708 | 0.5465 | 0.7332 | 0.7267 | 0.8013 | 0.5971 | 0.7281 |
| Thalamus | 0.8134 | 0.8969 | 0.5608 | 0.7813 | 0.7265 | 0.7958 | 0.5825 | 0.7046 |
| Frontal pole | 0.8107 | 0.8951 | 0.5698 | 0.7368 | 0.6920 | 0.7580 | 0.5802 | 0.6618 |
| Lateral parietal lobe | 0.7427 | 0.8174 | 0.6285 | 0.7343 | 0.6863 | 0.7562 | 0.6598 | 0.7183 |
| Basalganglia | 0.7696 | 0.8515 | 0.5513 | 0.7661 | 0.6814 | 0.7502 | 0.4778 | 0.6406 |
| Frontal gyri | 0.7473 | 0.8280 | 0.5494 | 0.6741 | 0.6822 | 0.7490 | 0.5962 | 0.6360 |
| Fusiforme gyri | 0.7383 | 0.8193 | 0.6501 | 0.7496 | 0.6789 | 0.7454 | 0.6140 | 0.7505 |
| Paracentral gyri | 0.7707 | 0.8523 | 0.5537 | 0.7296 | 0.6785 | 0.7453 | 0.6281 | 0.6902 |
| Medial frontal lobe | 0.7816 | 0.8646 | 0.5445 | 0.7586 | 0.6689 | 0.7338 | 0.6211 | 0.6993 |
| Medial parietal lobe | 0.7330 | 0.8118 | 0.5452 | 0.6523 | 0.6565 | 0.7148 | 0.5940 | 0.6556 |
| Temporal pole | 0.6979 | 0.7703 | 0.4810 | 0.6548 | 0.6494 | 0.7052 | 0.6036 | 0.6402 |
| Medial temporal lobe | 0.6797 | 0.7395 | 0.5834 | 0.7318 | 0.6463 | 0.7003 | 0.6060 | 0.7165 |
| Insula | 0.7424 | 0.8216 | 0.6262 | 0.7436 | 0.6297 | 0.6757 | 0.5799 | 0.6254 |
| Brainstem | 0.6592 | 0.7199 | 0.5632 | 0.5974 | 0.6177 | 0.6661 | 0.4700 | 0.5554 |
| **Whole brain** | **0.9459** | **0.9880** | **0.6983** | **0.9190** | **0.8978** | **0.9620** | **0.8311** | **0.9270** |

**Supplementary Table 2:** Accuracies and AUC for both models trained on the **volume-matched SHIP** data set when only **certain ROIs are visible** in the images.

| **ROI** | **LogReg** | | | | **BraiNN** | | | |
|---|---|---|---|---|---|---|---|---|
| | **SHIP** | | **HCP** | | **SHIP** | | **HCP** | |
| | **accuracy** | **AUC** | **accuracy** | **AUC** | **accuracy** | **AUC ↓** | **accuracy** | **AUC** |
| Cerebellum | 0.8655 | 0.9431 | 0.4830 | 0.8235 | 0.7700 | 0.8495 | 0.6948 | 0.7592 |
| Temporal gyri | 0.7632 | 0.8506 | 0.6059 | 0.7480 | 0.7364 | 0.8081 | 0.6123 | 0.7143 |
| Occipital lobe | 0.7640 | 0.8491 | 0.5452 | 0.6254 | 0.7259 | 0.8006 | 0.5829 | 0.6688 |
| Thalamus | 0.7648 | 0.8517 | 0.5938 | 0.7397 | 0.6948 | 0.7697 | 0.5334 | 0.6692 |
| Frontal pole | 0.7907 | 0.8802 | 0.5447 | 0.5844 | 0.6896 | 0.7686 | 0.4917 | 0.5075 |
| Amygdala Hippocampus | 0.7289 | 0.8055 | 0.6037 | 0.7512 | 0.6943 | 0.7582 | 0.5947 | 0.7294 |
| Frontal gyri | 0.7137 | 0.7867 | 0.5462 | 0.5962 | 0.6895 | 0.7547 | 0.5521 | 0.5750 |
| Medial temporal lobe | 0.6780 | 0.7397 | 0.5553 | 0.6172 | 0.6559 | 0.7186 | 0.5556 | 0.6231 |
| Paracentral gyri | 0.7272 | 0.8002 | 0.5629 | 0.6287 | 0.6584 | 0.7156 | 0.5850 | 0.6540 |
| Medial frontal lobe | 0.7287 | 0.8064 | 0.5460 | 0.5911 | 0.6463 | 0.7099 | 0.5496 | 0.5427 |
| Lateral parietal lobe | 0.6788 | 0.7453 | 0.5709 | 0.6758 | 0.6498 | 0.7031 | 0.6242 | 0.6717 |
| Fusiforme gyri | 0.6694 | 0.7386 | 0.5944 | 0.6357 | 0.6419 | 0.6980 | 0.5709 | 0.6356 |
| Basalganglia | 0.7091 | 0.7879 | 0.4719 | 0.6464 | 0.6144 | 0.6678 | 0.5217 | 0.5790 |
| Insula | 0.6972 | 0.7733 | 0.5518 | 0.5899 | 0.5893 | 0.6589 | 0.5261 | 0.5018 |
| Medial parietal lobe | 0.6822 | 0.7491 | 0.5442 | 0.5451 | 0.6111 | 0.6569 | 0.5384 | 0.5262 |
| Temporal pole | 0.6653 | 0.7261 | 0.5316 | 0.6151 | 0.6055 | 0.6559 | 0.5778 | 0.6115 |
| Brainstem | 0.6167 | 0.6741 | 0.5197 | 0.5212 | 0.5837 | 0.6204 | 0.4989 | 0.5281 |
| **Whole brain** | **0.9091** | **0.9690** | **0.7145** | **0.8290** | **0.8615** | **0.9300** | **0.7334** | **0.8430** |

**Supplementary Table 3:** Accuracies and AUC for both models trained on the **complete SHIP** data set when certain **ROIs are not visible** in the images.

| ROI | LogReg | | | | BraiNN | | | |
|---|---|---|---|---|---|---|---|---|
| | SHIP | | HCP | | SHIP | | HCP | |
| | **accuracy** | **AUC** | **accuracy** | **AUC** | **accuracy** | **AUC ↓** | **accuracy** | **AUC** |
| Fusiforme gyri | 0.9425 | 0.9871 | 0.6005 | 0.9390 | 0.9018 | 0.9649 | 0.8363 | 0.9267 |
| Medial temporal lobe | 0.9401 | 0.9871 | 0.6210 | 0.9369 | 0.9014 | 0.9646 | 0.8342 | 0.9250 |
| Medial frontal lobe | 0.9429 | 0.9873 | 0.6125 | 0.9366 | 0.8986 | 0.9640 | 0.8271 | 0.9246 |
| Insula | 0.9443 | 0.9880 | 0.5969 | 0.9313 | 0.8966 | 0.9632 | 0.8285 | 0.9244 |
| Amygdala Hippocampus | 0.9446 | 0.9878 | 0.6072 | 0.9465 | 0.8959 | 0.9630 | 0.8394 | 0.9260 |
| Temporal pole | 0.9414 | 0.9873 | 0.6049 | 0.9361 | 0.8991 | 0.9628 | 0.8280 | 0.9276 |
| Paracentral gyri | 0.9446 | 0.9873 | 0.6046 | 0.9373 | 0.8987 | 0.9628 | 0.8351 | 0.9266 |
| Medial parietal lobe | 0.9457 | 0.9881 | 0.6391 | 0.9492 | 0.8996 | 0.9625 | 0.8362 | 0.9272 |
| Frontal pole | 0.9410 | 0.9876 | 0.6073 | 0.9419 | 0.9001 | 0.9623 | 0.8369 | 0.9269 |
| Lateral parietal lobe | 0.9469 | 0.9881 | 0.6073 | 0.9399 | 0.8975 | 0.9619 | 0.8403 | 0.9282 |
| Frontal gyri | 0.9451 | 0.9888 | 0.6059 | 0.9457 | 0.8985 | 0.9618 | 0.8018 | 0.9286 |
| Basalganglia | 0.9436 | 0.9878 | 0.6020 | 0.9446 | 0.8975 | 0.9617 | 0.8349 | 0.9256 |
| Brainstem | 0.9438 | 0.9881 | 0.6072 | 0.9383 | 0.8990 | 0.9614 | 0.8362 | 0.9262 |
| Occipital lobe | 0.9430 | 0.9870 | 0.6309 | 0.9544 | 0.8982 | 0.9608 | 0.8408 | 0.9255 |
| Thalamus | 0.9426 | 0.9871 | 0.5771 | 0.9341 | 0.8939 | 0.9597 | 0.7878 | 0.9215 |
| Temporal gyri | 0.9463 | 0.9882 | 0.6055 | 0.9396 | 0.8915 | 0.9590 | 0.8371 | 0.9243 |
| Cerebellum | 0.9225 | 0.9784 | 0.6109 | 0.9378 | 0.8827 | 0.9521 | 0.8154 | 0.9051 |
| **Whole brain** | **0.9459** | **0.9880** | **0.6983** | **0.9190** | **0.8978** | **0.9620** | **0.8311** | **0.9270** |

**Supplementary Table 4:** Accuracies and AUC for both models trained on the **volume-matched SHIP** data set when certain **ROIs are not visible** in the images.

| **ROI** | **LogReg** | | | | **BraiNN** | | | |
|---|---|---|---|---|---|---|---|---|
| | **SHIP** | | **HCP** | | **SHIP** | | **HCP** | |
| | **accuracy** | **AUC** | **accuracy** | **AUC** | **accuracy** | **AUC ↓** | **accuracy** | **AUC** |
| Fusiforme gyri | 0.9042 | 0.9669 | 0.6976 | 0.8245 | 0.8633 | 0.9329 | 0.7166 | 0.8388 |
| Medial parietal lobe | 0.9050 | 0.9687 | 0.7266 | 0.8291 | 0.8623 | 0.9323 | 0.7109 | 0.8347 |
| Paracentral gyri | 0.9094 | 0.9715 | 0.6920 | 0.8380 | 0.8630 | 0.9312 | 0.7057 | 0.8355 |
| Frontal gyri | 0.9075 | 0.9713 | 0.7279 | 0.8472 | 0.8641 | 0.9306 | 0.7018 | 0.8378 |
| Temporal gyri | 0.9086 | 0.9706 | 0.7140 | 0.8184 | 0.8549 | 0.9282 | 0.7128 | 0.8277 |
| Lateral parietal lobe | 0.9039 | 0.9685 | 0.7076 | 0.8295 | 0.8651 | 0.9281 | 0.7233 | 0.8405 |
| Thalamus | 0.9135 | 0.9705 | 0.7067 | 0.8244 | 0.8611 | 0.9281 | 0.6735 | 0.8140 |
| Medial temporal lobe | 0.9005 | 0.9656 | 0.7107 | 0.8253 | 0.8537 | 0.9280 | 0.7108 | 0.8301 |
| Occipital lobe | 0.9111 | 0.9696 | 0.7394 | 0.8377 | 0.8543 | 0.9255 | 0.7174 | 0.8326 |
| Amygdala Hippocampus | 0.9038 | 0.9687 | 0.6983 | 0.8250 | 0.8526 | 0.9251 | 0.7168 | 0.8282 |
| Brainstem | 0.9078 | 0.9688 | 0.7143 | 0.8284 | 0.8552 | 0.9242 | 0.7015 | 0.8294 |
| Medial frontal lobe | 0.9050 | 0.9703 | 0.7275 | 0.8290 | 0.8598 | 0.9240 | 0.7128 | 0.8313 |
| Frontal pole | 0.9157 | 0.9724 | 0.7302 | 0.8384 | 0.8487 | 0.9214 | 0.7212 | 0.8235 |
| Insula | 0.9014 | 0.9671 | 0.7042 | 0.8333 | 0.8430 | 0.9213 | 0.7056 | 0.8326 |
| Temporal pole | 0.9090 | 0.9687 | 0.6959 | 0.8333 | 0.8496 | 0.9201 | 0.7117 | 0.8297 |
| Basalganglia | 0.9072 | 0.9669 | 0.6905 | 0.8307 | 0.8455 | 0.9174 | 0.7077 | 0.8193 |
| Cerebellum | 0.8798 | 0.9497 | 0.6494 | 0.7931 | 0.8422 | 0.9170 | 0.7129 | 0.8158 |
| **Whole brain** | **0.9091** | **0.9690** | **0.7145** | **0.8290** | **0.8615** | **0.9300** | **0.7334** | **0.8430** |
